# Performance and impact of disposable and reusable respirators for healthcare workers during pandemic respiratory disease: a rapid evidence review

**DOI:** 10.1101/2020.05.21.20108233

**Authors:** Chris Burton, Briana Coles, Anil Adisesh, Simon Smith, Elaine Toomey, Xin Hui Chan, Lawrence Ross, Trisha Greenhalgh

**Affiliations:** Academic Unit of Primary Medical Care, University of Sheffield; Diabetes Research Centre, University of Leicester, UK; Department of Medicine, Division of Occupational Medicine, University of Toronto; Division of Occupational Medicine, St. Michael’s Hospital, Unity Health, Toronto; Chair, Canadian Standards Biological Aerosols Working Group; School of Allied Health, University of Limerick, Limerick, Ireland; Centre for Tropical Medicine and Global Health, Nuffield Department of Medicine, University of Oxford; Department of Infectious Disease, Children’s Hospital of Los Angeles; Nuffield Department of Primary Care Health Sciences, University of Oxford

**Author notes:** **Address for correspondence:** Professor Chris Burton, Academic Unit of Primary Medical Care, University of Sheffield Herries Rd, Sheffield, S5 7AU, UK.

## Abstract

**Objectives:** In the context of the Covid-19 pandemic, to identify the range of filtering respirators that can be used in patient care and synthesise evidence to guide the selection and use of different respirator types.

**Design:** Comparative analysis of international standards for filtering respirators and rapid review of their performance and impact in healthcare.

**Data sources:** Websites of international standards organisations, Medline and EMBASE (final search 11^th^ May 2020), with hand-searching of references and citations.

**Study selection:** Guided by the SPIDER tool, we included studies whose sample was healthcare workers (including students). The phenomenon of interest was respirators, including disposable and reusable types. Study designs including cross-sectional, observational cohort, simulation, interview and focus group. Evaluation approaches included test of respirator performance, test of clinician performance or adherence, self-reported comfort and impact, and perceptions of use. Research types included quantitative, qualitative and mixed methods. We excluded studies comparing the effectiveness of respirators with other forms of protective equipment.

**Data extraction, analysis and synthesis:** Two reviewers extracted data using a template. Suitability for inclusion in the analysis was judged by two reviewers. We synthesised standards by tabulating data according to key criteria. For the empirical studies, we coded data thematically followed by narrative synthesis.

**Results:** We included relevant standards from 8 authorities across Europe, North and South America, Asia and Australasia. 39 research studies met our inclusion criteria. There were no instances of comparable publications suitable for quantitative comparison. There were four main findings. First, international standards for respirators apply across workplace settings and are broadly comparable across jurisdictions. Second, effective and safe respirator use depends on proper fitting and fit-testing. Third, all respirator types carry a burden to the user of discomfort and interference with communication which may limit their safe use over long periods; studies suggest that they have little impact on specific clinical skills in the short term but there is limited evidence on the impact of prolonged wearing. Finally, some clinical activities, particularly chest compressions, reduce the performance of filtering facepiece respirators.

**Conclusion:** A wide range of respirator types and models is available for use in patient care during respiratory pandemics. Careful consideration of performance and impact of respirators is needed to maximise protection of healthcare workers and minimise disruption to the delivery of care.

## Background

The global Covid-19 pandemic has increased demand worldwide for respirators to use in direct patient care^1-3^. This includes both disposable devices (such as filtering facepiece respirators) and reusable ones (such as elastomeric and powered air-purifying respirators). Staff previously unfamiliar with these devices are now required or advised to use them. Shortages of supply have also led to consideration of “repurposing” respirators from other industries for healthcare use^4^.

This review is designed to inform front-line healthcare professionals, occupational health advisers and policymakers about the performance and impact of respirators, particularly in the context of the Covid-19 pandemic. We have focused on the performance and impact of different types of respirator in relation to clinical use. By ‘performance’, we refer to the level of protection provided by respirators (for example in laboratory studies of filtering capability or in practical use), and by ‘impact’ we refer to the effects on clinical activities of wearing one. The comparative effectiveness of respirators against other equipment, and guidelines for when respirators should be used, were beyond the scope of this review.

### What is a respirator?

A filtering respirator is a personally-worn item of protective equipment which removes hazardous materials from inhaled air. It is designed to be used in conjunction with other protective equipment as an “ensemble”.^5 6^ These respirators work by filtering air either by negative pressure (the work of inspiration pulls air through a filter) or positive pressure (a blower draws air through a filter and feeds that to the user). Respirators which use negative pressure require an airtight seal against the user’s face to ensure that inspired air passes through, rather than around, the filter. Respirators which use a blower are less dependent on a tight seal and can include a loose-fitting hood.

In healthcare, respirator filters – either in the mask itself or in a filter housing – are used to filter aerosols containing infectious agents. Filters comprise a multi-layered fibrous web; most modern filters also incorporate an electrostatically charged layer to enhance capture of very small particles. This allows the web to be more open and afford more comfortable breathing while still protecting the user. For necessary protection, both an adequate filter and an adequate fit to the wearer are needed. Respirators are available in a wide range of types and designs. Broadly, there are three types relevant to healthcare: filtering facepiece respirators (FFR, including the FFP2 and N95 mask); elastomeric facepiece respirators (EFR); and powered air-purifying respirators (PAPR). Box 1 provides further detail on these different kinds.

##### Box 1: Different kinds of respirator

###### Filtering Facepiece Respirators (FFRs)

Filtering facepiece respirators are made of moulded filter material, shaped to form a tight seal with the wearer’s face, such that inspired air must pass through the filter layers. They differ from simple masks (including fluid-resistant surgical masks) which permit airflow around the mask. Most filtering facepiece respirators involve the user breathing in and out through the filter, though some models incorporate a valve to allow exhaled air to vent directly. The level of user protection depends on the integrity of the seal to the face. Filtering facepiece respirators are generally discarded after hours or a day of use, but shortages in emergency may lead to their re-use.

###### Elastomeric Facepiece Respirators (EFRs)

Elastomeric facepiece respirators generally incorporate a plastic facepiece with an elastomeric (often silicone rubber) seal against the face. The most common respirator in this category in healthcare has a half-facepiece so requires additional eye protection, but full-face versions are also used. An exhalation valve is always present and there are attachments for one or two filters. In most cases, filters are replaceable, often with a choice of types appropriate to the hazard. The facepiece is designed to be decontaminated and used repeatedly. Some models include a speech transmission diaphragm to assist communication by wearer.

###### Powered Air Purifying Respirators (PAPRs)

Powered air purifying respirators incorporate a piece of headgear which receives air, drawn through a filter by a motor-driven fan. Filters are fitted on to the blower unit appropriate to the hazard. PAPRs used in healthcare typically have a body-worn blower connected to a headpiece by a hose. The headgear can either be a loose-fitting hood or a tight fitting (sealed) mask. The loose-fitting hood type does not need a seal because the positive pressure ensures a constant outflow from the hood. One advantage of PAPRs is that they remove the effort of breathing against the resistance of filters, and so reduce the wearer’s physiological burden. They can also accommodate facial conformities where a face fit seal has been unsuccessful including for users with beards. Blowers generally employ rechargeable batteries (though for emergency stockage primary cells may be available), so a battery maintenance programme is necessary, as is an air flowrate check before use.

### Fit testing

The effectiveness of a respirator depends on two things: its filtration performance and its effective use by the wearer to avoid inhaling unfiltered air. It is necessary to carry out a medical evaluation to ensure fitness to use a respirator, and a workplace risk assessment to match the expected exposure. Part of this assessment is a formal fit test which ensures an adequate seal to the size and shape of the face of the user. Fit testing can be either qualitative (awareness of a sweet or bitter aerosol) or quantitative (measurement of aerosol ingress) with evaluation while various head and body movements and breathing and speaking exercises are performed. Fit testing requires trained personnel and specific equipment. Loose-fitting powered air-purifying respirators do not require fit testing. Once a user’s respirator fit has been tested, they are trained to perform a face seal check – typically by breathing in or out sharply to check for leakage around the respirator - each time a fit-tested facepiece is worn and before entering a hazardous environment.

## Review Question

### Overall question

What is the range of disposable and reusable respirators that can be used for infection control purposes in patient care, what evidence guides the selection and use or respirator type, and how can this knowledge be used to address the needs of the Covid-19 and future respiratory pandemics?

### Specific Questions

1. What standards currently exist for respirators in healthcare and non-healthcare settings and how do these standards compare?
2. How well do respirators perform in clinical settings in terms of fit, either initially or during clinical activities?
3. How do healthcare workers and organisations use and perceive different forms of respirator in practice?
4. What are the impacts on clinicians and their performance of using different forms of respirators in patient care?

### Context and scope

We aimed to address the question in the context of clinical care for patients with proven or likely Covid-19 in high risk settings where there is a substantial risk to professionals from the presence of virus-containing aerosols. A rapid review to create a taxonomy of aerosol-generating medical procedures and scenarios is being carried out in parallel with this review and will be published separately.

This review aims to summarise the evidence for frontline clinicians, occupational health leads and policymakers. It recognises that in times of extreme demand for respiratory protective equipment, such as the Covid-19 pandemic, it is reasonable to ensure that the full range of respiratory protection options is considered. We aimed to review the evidence from both a selection of formal standards and published clinical research in order to support users to make informed decisions and choices.

## Methods

### Review type

This rapid review was informed by the Cochrane Rapid Reviews Interim Guidance produced to guide the rapid generation of evidence syntheses in the Covid-19 pandemic.^7^ The protocol was made available at the Open Science Framework on 3^rd^ May 2020 and finalised on 11th May^8^ while data extraction was in progress but before it was completed.

### Searches and identifying literature

#### Identification and comparison of standards

We searched documentation and websites of standards organisations from Europe, North America (Canada, USA, Mexico), Australia, and Asia (China, Japan and Korea) for information relating to standards for filtering respirators. This was informed and supplemented by in-depth specialist knowledge of regulatory processes and standards for respirators of one of the authors (SS).

We compared standards by tabulating the extracted data according to key criteria. Fields for the framework include geopolitical region; standard reference and year; respiratory protective equipment classification within the standard; test agent; and maximum permitted inward leakage.

#### Performance and impact in the context of healthcare

We conducted a systematic search to identify studies examining the performance of respiratory protective equipment in healthcare contexts. We took a mixed methods approach, which allowed us to include data from diverse study types including survey, direct observation of practice, observation and measurement at rest or in simulated clinical activity and qualitative studies relating to perceptions about the use of respirators.

We searched Medline and EMBASE for papers published before 1^st^ May 2020 (updated 13^th^ May 2020). This was supplemented by prior expert knowledge of one of the team (SS) from working in respirator manufacture and contribution to Canadian and other international standards and by handsearching of references and citations from key papers^9^. The search was designed to be sufficiently inclusive to address research questions 2 and 3. Eligibility criteria were framed using the SPIDER tool:^10^

- Sample – healthcare workers or student healthcare workers
- Phenomenon of Interest – respirators: including disposable filtering facepiece and reusable (elastomeric filtering facepiece and powered air-purifying) types
- Design – wide range of designs including cross-sectional, cohort observation, simulation and interview or focus group
- Evaluation – either (a) test of respirator performance, or (b) test of clinician performance or adherence, or (c) self-reported comfort and impact, or (d) perceptions of use.
- Research types: quantitative, qualitative or mixed-method.

Detailed search terms are listed in appendix 1.

Titles and abstracts from the search results were screened by one reviewer (CB). A second reviewer (BC) reviewed a randomly selected 20% of titles and abstracts. The first reviewer then screened all full texts for inclusion and the second checked those which had been excluded. For practical purposes, the search strategy was designed to be moderately restrictive (returning between 100 and 500 titles). We limited data extraction to peer-reviewed papers or full-text pre-prints in English.

### Data extraction & synthesis

Data was extracted from identified papers by CB and BC using a template in Google Forms feeding to a spreadsheet. The template linked papers to specific research questions and sub questions, although papers could be included in addressing more than one research question.

From the extracted data, two authors (CB and BC) created a table of summary characteristics and key findings. We conducted a narrative synthesis of the study findings in which similar studies were grouped by themes. No meta-analysis was carried out as insufficient studies reported a comparable quantitative measure. Finally, a summary of evidence table was developed by two authors (CB and BC) which summarised the main findings according to key themes and the types of studies contributing to each theme.

## Results

### Search results

The search of Medline and Embase returned 394 papers and a further 26 were identified by following references and citations and from personal archives. More detail is provided in the PRISMA diagram (Figure 1).

**Figure 1:**
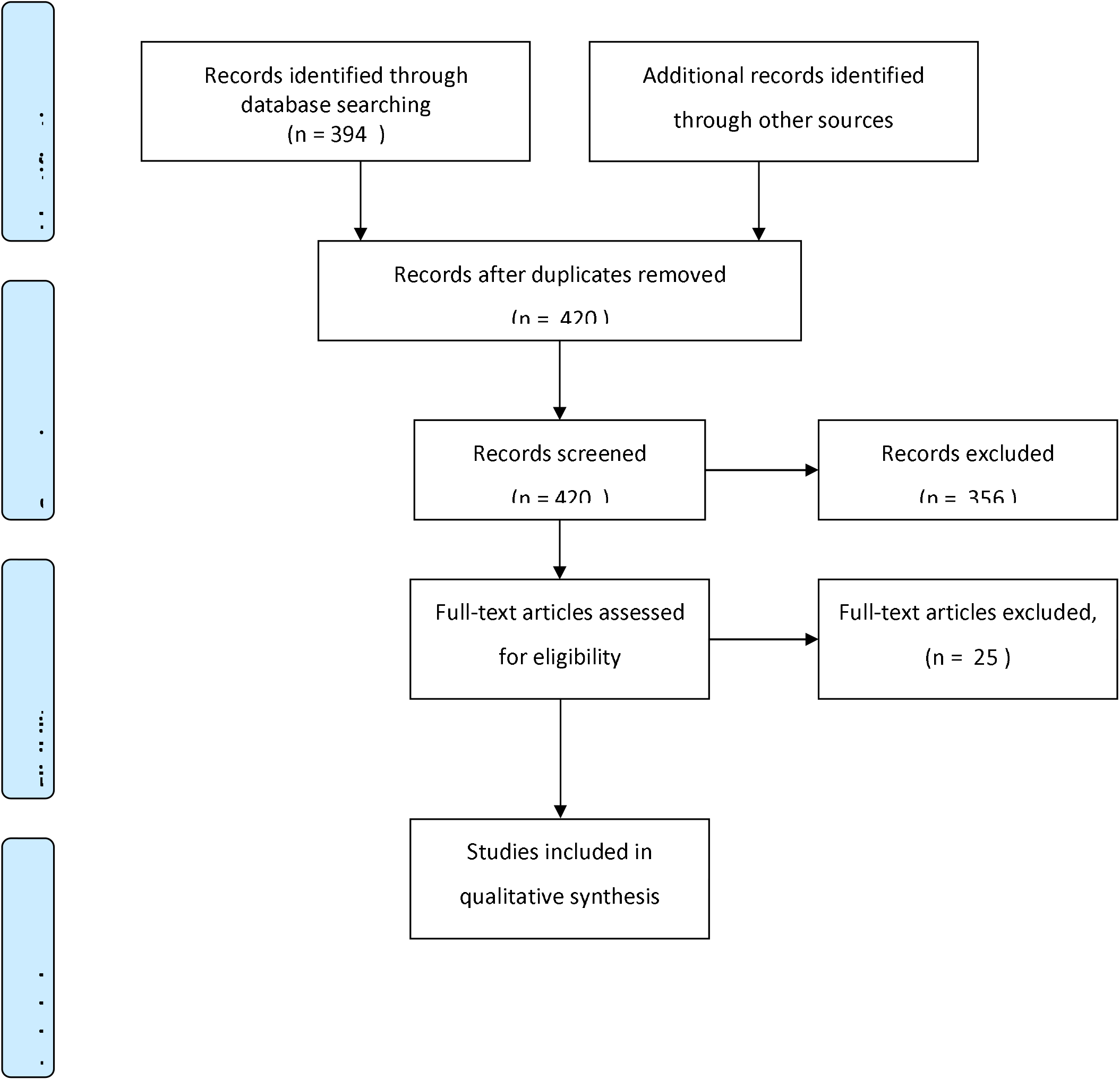
PRISMA 2009 Flow Diagram.

### Review of performance standards and approvals

Performance standards for filtering respirators are set by national and international standards organisations. Standards relate both to the performance of devices and to their selection and use in the workplace. **Error! Reference source not found**. lists major standards organisations, the countries in which the standards apply, and the main standards relating to respirator performance, selection and use.

Performance standards for respirators include the ability of the device to filter particles from inspired air. Filter penetration is typically tested with an aerosol of sodium chloride or aerosols of paraffin oil or dioctyl phthalate. These substances have similar penetration properties to biological aerosols encountered in healthcare settings. Standards also include measures of resistance to penetration by airborne materials, of resistance to breathing (both inspiration and expiration) and maximum permitted CO_2_ build-up.

**Error! Reference source not found**. lists the performance of widely-recognised filtering respirator classifications. This includes standards for filtering facepiece respirators (e.g. FFP2 and FFP3 in Europe, N95 and P100 in North America and P2 and P3 in Australia). While these standards are not identical, there are strong similarities between standards (e.g. N95 classification is comparable to FFP2). Similar standards apply to the filters for use with other respirators such as elastomeric facepiece respirator and powered air-purifying respirators. Some N95 or equivalent respirators have additionally been cleared by regulatory authorities to meet surgical mask fluid penetration requirements. Even if not formally cleared, filtering facepiece respirators generally offer useful fluid resistance, and with types for which approval testing includes oil-based aerosols, this is likely to be high, but in all cases manufacturers’ direction should be followed. There have been recommendations to wear a surgical mask over a filtering facepiece respirator^11^, however this does not increase respiratory protection and does increase the burden to the wearer.^12 13^ We did not consider extended use or reuse of respirators in this review as that topic is the subject of a separate review.^14^

The standards reported in **Error! Reference source not found**. and **Error! Reference source not found**. are not specific to healthcare. Therefore, a respirator (either disposable or reusable) may be used in a range of different settings, providing that the standards it meets are those applicable in the new setting. All standards documents are explicit that supplying a respirator is only one part of a respiratory protection programme and that ensuring adequate fit and safe use is essential.

### Review of research literature on performance and impact

We identified 39 eligible original publications, no relevant systematic reviews and one narrative review from 2015 which did not provide a systematic search strategy.^15^ We also identified a recent edited book on elastomeric respirators.^16^ We grouped findings into seven themes: assessing respirator fit; the effect of clinical activities on respirator fit; respirator use in practice and the effects of training; impact of respirator use on clinical performance; impact on communication; impact on the user; and adoption of respirator use by individuals and organisations.

Table 3 and Table 4 summarise the primary studies identified in our search.

**Table 1.** Standards authorities for respiratory protective devices and major relevant standards.

| Organisation | Recognised in | Respirator performance standards<br>(includes requirements, testing & marking)<br>Latest revision year indicated |  | Selection, use and care standards (or nearest equivalent)<br>(includes user testing and appropriate use) |  |
| --- | --- | --- | --- | --- | --- |
|  |  | Standard | Description | Standard | Description |
| Australia/New Zealand Standards (AS/NZS) | Australia & New Zealand | AS/NZS 1716 (2012) | Respiratory Protective Devices | AS/NZS 1715 (2012) | Selection, use and maintenance of respiratory protective equipment |
| Brazil Associação Brasileira de Normas Técnicas (ABNT) | Brazil | ABNT NBR 13698 (2011) | Respiratory protective devices - Filtering half mask to protect against particles | ABNT NBR 12543 (2017) | Respiratory protective devices - Terminology |
| Canadian Standards Association (CSA) | Canada |  |  | CSA Z94.4 (2018) | Selection, use and care of respirators |
| Standardization Administration of China | China | GB 2626 (2019) | Non-powered air-purifying particle respirators | GB/T 18664 (2002) | Selection, use and maintenance of Respiratory protective equipment |
|  |  | GB 30864 (2014) | Powered air-purifying respirators |  |  |
| European Committee for Standardization (CEN) | UK, European Union, European Free-Trade Association, Russia, South Africa | EN 149 (2009) | Filtering facepiece | EN 132 (1999) | Definitions of terms & pictograms |
|  |  | EN 136 & EN 140 (1998) | Elastomeric facepiece | EN 529 (2005) | Recommendations for selection, use, care and maintenance |
|  |  | EN 12941 (2008) | Loose fitting PAPR |  |  |
|  |  | EN 12942 (2008) | Tight-fitting PAPR |  |  |
|  |  | EN 143 (2000) | Filters for respirators |  |  |
| Japanese Industrial Standards Committees (JIS)<sup>1</sup> | Japan | JIS T 8151 (2018) | Particulate respirators | JIS T 8150 (2006) | Guidance for selection, use and maintenance of respiratory protective devices |
|  |  | JIS T 8157 (2018) | Powered air purifying respirator for particulate matter |  |  |
| Japan Ministry of Health, Labour and Welfare (JMHLW) |  | Notification 214-2018 | Standard for Dust Mask |  |  |
| Korean Agency for Technology and Standards (KATS)<sup>2</sup> | Korea | KS M 6673 (2008) | Dust respirators | KS P 1101 (2010) | Guidance for selection, use and maintenance of respiratory protective devices |
|  |  | KS M 6764 (2009) | Filter for dust respirators |  |  |
|  |  | KS P 8416 (2008) | Dust respirators for fine particles |  |  |
|  |  | KS P 8417 (2008) | Powered air purifying respirators |  |  |
| Korean Ministry of Employment and Labour (KMOEL) |  | KMOEL Notification 2017-64 (2017) | Dust respirators |  |  |
| Mexican Norma Oficial Mexicana (NOM) | Mexico | NOM-116-STPS-2009 | Particulate FFP and replaceable filters | Annex to NOM-116-STPS-2009 | Guide for selection of air purifying respirators for hazardous dusts |
| U.S. National Institute for Occupational Safety & Health (NIOSH) | USA, Canada<sup>1</sup> | 42 CFR 84 (1995) | All types of respiratory protective device | 29 CFR 1910.134 (1998) (USA only) | Respiratory Protection |

<sup>1</sup> In Japan, JIS standards are not mandatory, while JMHLW notifications are mandatory
<sup>2</sup> In Korea, KATS standards are not mandatory, while KMOEL notifications are mandatory

**Table 2.** Details of standards for filtering facepiece respirators and filters for reusable respirators.

| Domain | Respiratory Protective Equipment Classification (FFR and reusable filter) | Minimum efficiency of filter performance<sup>2</sup> |  | FFR Maximum total inward leakage | Tested for oil atmosphere<sup>3</sup> | FFR Maximum inhalation airflow resistance<sup>4</sup> |  | FFR Maximum exhalation airflow resistance |  | FFR/EFR Maximum CO<sub>2</sub> build-up |
| --- | --- | --- | --- | --- | --- | --- | --- | --- | --- | --- |
|  |  | Value | Flow (L/min) |  |  | Value (Pa) | Test Flow (L/min) | Value (Pa) | Test Flow (L/min) |  |
| Australia | P1 respirator / P1 filter | 80% | 95 | 22% | All types | 60/210 | 30/95 | 120 | 85 | 1% |
|  | P2 respirator / P2 filter | 94% |  | 8% |  | 70/240 | 30/95 |  |  |  |
|  | P3 respirator / P3 filter | 99% |  | 2% |  | 100/300 | 30/95 |  |  |  |
| Brazil | PFF1 S / PFF1 SL respirator / P1 filter | 80% | 95 | Not specified | Not S-types | 60/210 | 30/95 | 120 | 85 | 1% |
|  | PFF2 S / PFF2 SL respirator / P2 filter | 94% |  |  |  | 70/240 | 30/95 |  |  |  |
|  | PFF3 S / PFF3 SL respirator / P3 filter | 99% |  |  |  | 100/300 | 30/95 |  |  |  |
| China | KN95 / KR95 / KP95 | 95% | 85 | 8% | Not KN-types | 350 | 85 | 250 | 85 | 1% |
|  | KN99 / KR99 / KP99 | 99% |  |  |  |  |  |  |  |  |
|  | KN100 / KR100 / KP100 | 99.97% |  |  |  |  |  |  |  |  |
| Europe | FFP1 respirator / P1 filter | 80% | 95 | 22% | All types | 60/210 | 30/95 | 300 | 160 | 1% |
|  | FFP2 respirator / P2 filter | 94% |  | 8% |  | 70/240 | 30/95 |  |  |  |
|  | FFP3 respirator / P3 filter | 99% |  | 2% |  | 100/300 | 30/95 |  |  |  |
| Japan | DS1 / DL1 respirator / RS1 / RL1 filter | 80% | 85 | See footnote<sup>5</sup> | Not DS or RS types | 60/45 | 85 | 60/45<sup>6</sup> | 85 | 1% |
|  | DS2 / DL2 / RS2 / RL2 filter | 95% |  |  |  | 70/50 | 85 | 70/50 | 85 |  |
|  | DS3 / DL3 / RS3 / RL3 filter | 99.9% |  |  |  | 150/100 | 85 | 80/60 | 85 |  |
| Korea | KF80 (2nd Class) | 80% | 95 | 22% | All types | 60/210 | 30/95 | 300 | 160 | 1% |
|  | KF94 (1<sup>st</sup> Class) | 94% |  | 8% |  | 70/240 | 30/95 |  |  |  |
|  | Special | 99.9% |  | 2% |  | 100/300 | 30/95 |  |  |  |
| Mexico | N90 / R90 / P90 | 90% | 85 | Not specified | Not N types | 343 | 85 | 245 | 85 | None |
|  | N95 / R95 / P95 | 95% |  |  |  |  |  |  |  |  |
|  | N100 / R100 / P100 | 99.97% |  |  |  |  |  |  |  |  |
| USA & Canada | N95 / R95 / P95 | 95% | 85 | No requirement<sup>7</sup> | Not N types | 343 | 85 | 245 | 85 | None |
|  | N99 / R99 / P99 | 99% |  |  |  |  |  |  |  |  |
|  | N100 / R100 / P100 | 99.97% |  |  |  |  |  |  |  |  |

FFR: Filtering facepiece respirator, EFR Elastomeric facepiece respirator
<sup>1</sup> In Canada, there are multiple jurisdictions: NIOSH approvals are generally accepted but those of other agencies may also be applicable in some jurisdictions
<sup>2</sup> Minimum efficiency at most penetrating particle size – typically 0.2-0.3 micron mass median diameter
<sup>3</sup> Testing performance in an oil atmosphere is an indicator of additional fluid resistance, the clinical relevance of this is uncertain.
<sup>4</sup> Dual values indicate testing at two flow rates, single values indicate testing at one flow rate
<sup>5</sup> Inward Leakage measured, included in user Instructions
<sup>6</sup> First value is for FFP without exhalation valves, second value for FFP with exhalation valves.
<sup>7</sup> No requirement in this standard, though max. 10% in practice

**Table 3.** Study characteristics - performance of respirators and healthcare workers using them.

| Study | N | Study design | Respirators | Type of activity | Comparator | Primary Outcome | Findings |
| --- | --- | --- | --- | --- | --- | --- | --- |
| Danyluk 2011<sup>18</sup> | 784 | Cross sectional testing | Filtering facepiece | - | QNFT & QLFT | seal check vs QNFT | 643 respirator naive passed seal check with appropriate device: 92/643 failed QLFT, 158/643 failed QNFT. Results no different for experienced 30/137 & 41/137. Comparison of QNFT & QLFT in Hon 2016 |
| Derrick 2005<sup>19</sup> | 93 | Cross sectional testing | Mixed | - | QNFT | seal check vs QNFT | The user seal check wrongly indicated that the mask fitted on 18–31% of occasions, and wrongly indicated that it did not fit on 21–40% of occasions. (insufficient data for sensitivity and specificity) |
| Kim 2019<sup>31</sup> | 22 | Before after training | Filtering facepiece | - | QNFT | Fit factor | Fit factors, overall fit factor, and adequate protection rate were higher after training than before training for the 3 types of respirators (all $p < .05$). |
| Lam 2016<sup>17</sup> | 638 | Cross-sectional | Filtering facepiece | - | QNFT | seal check vs QNFT | LR for seal test close to 1; Sen 22-28% Spec 76-82% |
| Lee 2008<sup>33</sup> | 43 | Cohort, longitudinal | Filtering facepiece | - | QNFT | repeated fit tests | Training and fitting got 100% initial pass but slipped to 46% at 3 month follow up without further training (boosted by reminder and that provided better response at 14 months) |
| Lee 2017<sup>23</sup> | 25 | Crossover | Filtering facepiece | - | QNFT | Fit test | Fold type N95 good performance with 100% passing fit test for most actions, Cup and valve types <50% satisfactory fit |
| McMahon 2008<sup>20</sup> | 1271 | Cross-sectional testing | Filtering facepiece | - | QNFT | Fit test | 95% men and 85% women passed at first fitting. Almost all remainder eventually fitted. Essential to have range of respirators to ensure satisfactory fit |
| Sandaradura 2020<sup>22</sup> | 105 | Cross sectional testing | Filtering facepiece | - | QNFT | fit factor | Relative to those with no facial hair, the OR for respirator fit was 0.74 (95% CI 0.21-2.52) for light stubble, 0.45 (95% CI 0.12-1.57) for moderate to heavy stubble, 0.04 (95% CI 0-0.28) for full beard and 0.56 [95% CI 0.05-4.48] for other types of facial hair. |
| Wilkinson 2010<sup>21</sup> | 5024 | Cross sectional testing | Filtering facepiece | - | QNFT | Fit test | 4472/5024 (89%) got successful fit; 3707/4472 (83%) got fit first time |
| Winter 2010<sup>32</sup> | 50 | Cross sectional testing | Filtering facepiece | - | QLFT | Fit test | pre-training, first mask 9/50 passed; post training, first mask 20/50; post training best fitting 36/50 passed. |
| Hauge 2012<sup>30</sup> | 8 | Cohort testing |  | Simulated nursing activity | QNFT | Fit factor | all participants had good fit at baseline, all maintained FF>100 in activity |
| Hwang 2020<sup>24</sup> | 44 | Cohort simulation | Filtering facepiece | Simulated CPR | QNFT | fit factor during activity | Overall, 73% (n = 32) of the participants failed at least one of the three chest compression sessions |
| Kang 2018<sup>28</sup> | 26 | Cohort simulation | Filtering facepiece | Simulated intubation | QNFT | fit factor during activity | FF<100 using cup masks and direct laryngoscope in 25% of intubations. No problem with folding filtering facepiece or with video laryngoscope or laryngeal mask airway. |
| Park 2020<sup>27</sup> | 91 | Cohort simulation | PAPR | Simulated CPR | QNFT | Fit Factor during activity | All participants maintained FF > 100 throughout. High acceptability, but 20% reported difficulty hearing. |
| Shin 2017<sup>26</sup> | 30 | Crossover | Filtering facepiece | Simulated CPR | QNFT | fit factor during activity | Fit factor falls during chest compression, <50% protected with cup design FFR, >90% protected with fold type. |
| Sietsema 2018<sup>25</sup> | 15 | Cohort simulation | Filtering facepiece | Simulated nursing activity | QNFT | correlation fit factor pre & during activity | overall resting and simulated workplace factors were highly correlated (r=0.88, p < 0.001) |
| Suen 2017<sup>29</sup> | 120 | Cohort simulation | Filtering facepiece | Simulated nursing activity | QNFT | fit factor before and after activity | Fit factor fell after activity: in 40/120 post activity was below 100 |
| Beam 2018<sup>34</sup> | 24 | Observational | Filtering facepiece respirator | Actual | Standard | Adherence to standards | 10/24 incorrectly donned FFR; 10/24 adjusted while in room; only 2/24 did manual seal check before entry into room. |
| Mumma 2019<sup>36</sup> | 41 | Observational | PAPR | Actual | Standard | NA | Donning and doffing study: PAPR hood removal associated with some contamination risk |
| Nichol 2013<sup>35</sup> | 100 | Observational | Filtering facepiece | Simulated | Standard | NA | 44% passed 5/6 criteria. Lowest pass rates for seal check (24%) & not touching (only 40%). Critical care more likely to pass than emergency department (suggests familiarity) |

CI confidence interval; CPR: cardiopulmonary resuscitation; EFR Elastomeric facepiece respirator; FF: Fit factor; FFR: Filtering facepiece respirator; NA: not applicable; PAPR: powered air-purifying respirator; QNFT: quantitative fit test; QLFT: qualitative fit test

**Table 4.**
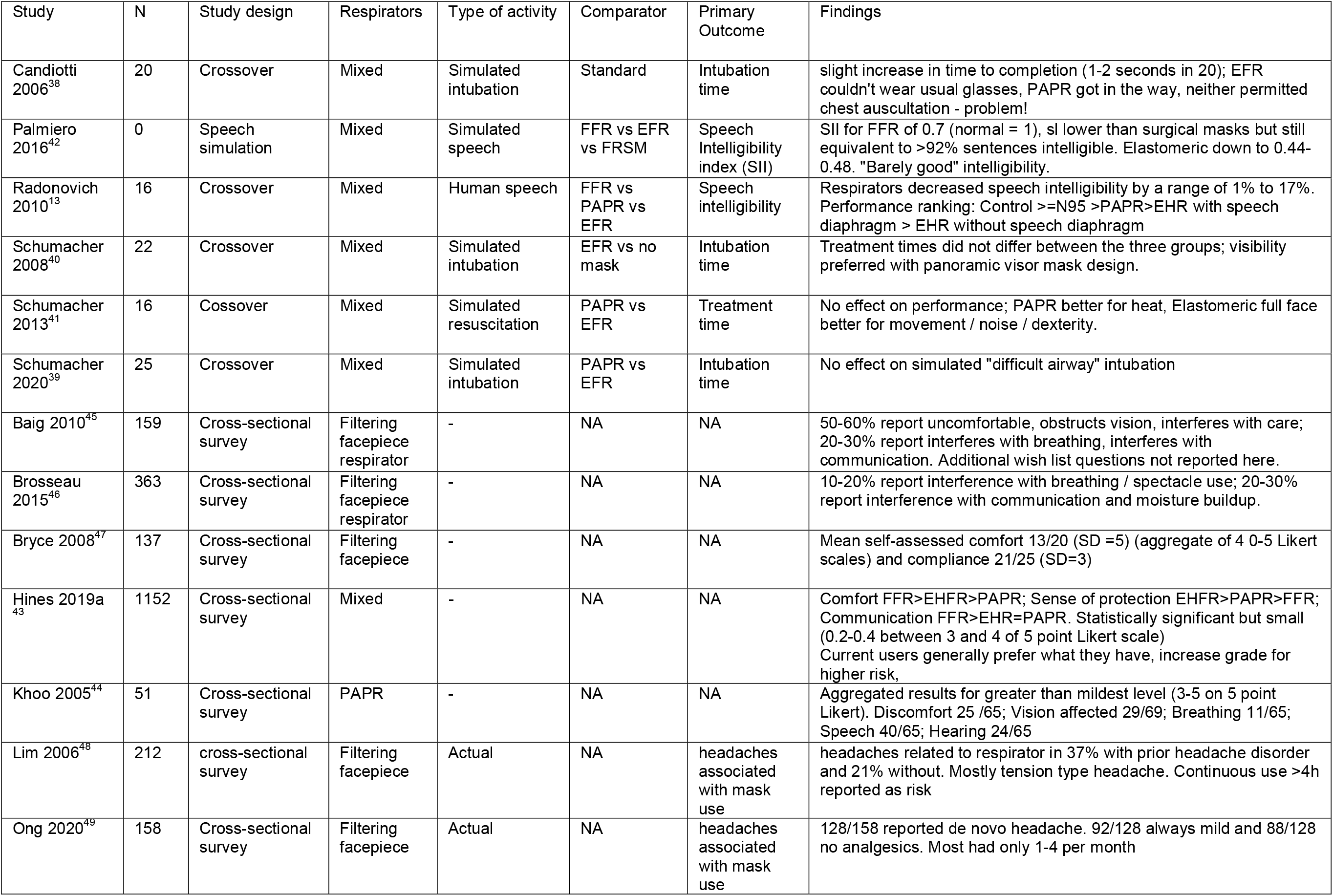
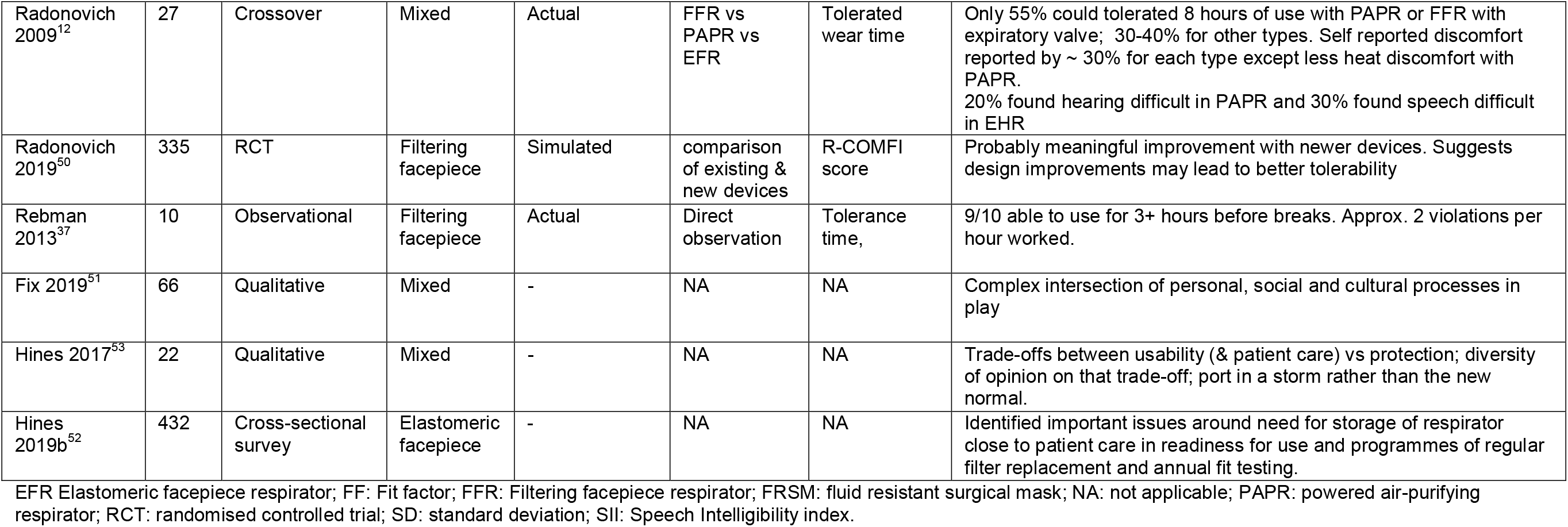
Study characteristics – impact of respirators on healthcare workers clinical activities and comfort.

| Study | N | Study design | Respirators | Type of activity | Comparator | Primary Outcome | Findings |
| --- | --- | --- | --- | --- | --- | --- | --- |
| Candiotti 2006<sup>38</sup> | 20 | Crossover | Mixed | Simulated intubation | Standard | Intubation time | slight increase in time to completion (1-2 seconds in 20); EFR couldn't wear usual glasses, PAPR got in the way, neither permitted chest auscultation - problem! |
| Palmiero 2016<sup>42</sup> | 0 | Speech simulation | Mixed | Simulated speech | FFR vs EFR vs FRSM | Speech Intelligibility index (SII) | SII for FFR of 0.7 (normal = 1), sl lower than surgical masks but still equivalent to >92% sentences intelligible. Elastomeric down to 0.44-0.48. "Barely good" intelligibility. |
| Radonovich 2010<sup>13</sup> | 16 | Crossover | Mixed | Human speech | FFR vs PAPR vs EFR | Speech intelligibility | Respirators decreased speech intelligibility by a range of 1% to 17%. Performance ranking: Control >=N95 >PAPR>EHR with speech diaphragm > EHR without speech diaphragm |
| Schumacher 2008<sup>40</sup> | 22 | Crossover | Mixed | Simulated intubation | EFR vs no mask | Intubation time | Treatment times did not differ between the three groups; visibility preferred with panoramic visor mask design. |
| Schumacher 2013<sup>41</sup> | 16 | Cosrossover | Mixed | Simulated resuscitation | PAPR vs EFR | Treatment time | No effect on performance; PAPR better for heat, Elastomeric full face better for movement / noise / dexterity. |
| Schumacher 2020<sup>39</sup> | 25 | Crossover | Mixed | Simulated intubation | PAPR vs EFR | Intubation time | No effect on simulated "difficult airway" intubation |
| Baig 2010<sup>45</sup> | 159 | Cross-sectional survey | Filtering facepiece respirator | - | NA | NA | 50-60% report uncomfortable, obstructs vision, interferes with care; 20-30% report interferes with breathing, interferes with communication. Additional wish list questions not reported here. |
| Brosseau 2015<sup>46</sup> | 363 | Cross-sectional survey | Filtering facepiece respirator | - | NA | NA | 10-20% report interference with breathing / spectacle use; 20-30% report interference with communication and moisture buildup. |
| Bryce 2008<sup>47</sup> | 137 | Cross-sectional survey | Filtering facepiece | - | NA | NA | Mean self-assessed comfort 13/20 (SD =5) (aggregate of 4 0-5 Likert scales) and compliance 21/25 (SD=3) |
| Hines 2019a<sup>43</sup> | 1152 | Cross-sectional survey | Mixed | - | NA | NA | Comfort FFR>EHFR>PAPR; Sense of protection EHFR>PAPR>FFR; Communication FFR>EHR=PAPR. Statistically significant but small (0.2-0.4 between 3 and 4 of 5 point Likert scale)<br>Current users generally prefer what they have, increase grade for higher risk, |
| Khoo 2005<sup>44</sup> | 51 | Cross-sectional survey | PAPR | - | NA | NA | Aggregated results for greater than mildest level (3-5 on 5 point Likert). Discomfort 25 /65; Vision affected 29/69; Breathing 11/65; Speech 40/65; Hearing 24/65 |
| Lim 2006<sup>48</sup> | 212 | cross-sectional survey | Filtering facepiece | Actual | NA | headaches associated with mask use | headaches related to respirator in 37% with prior headache disorder and 21% without. Mostly tension type headache. Continuous use >4h reported as risk |
| Ong 2020<sup>49</sup> | 158 | Cross-sectional survey | Filtering facepiece | Actual | NA | headaches associated with mask use | 128/158 reported de novo headache. 92/128 always mild and 88/128 no analgesics. Most had only 1-4 per month |
| Radonovich 2009<sup>12</sup> | 27 | Crossover | Mixed | Actual | FFR vs PAPR vs EFR | Tolerated wear time | Only 55% could tolerate 8 hours of use with PAPR or FFR with expiratory valve; 30-40% for other types. Self reported discomfort reported by ~ 30% for each type except less heat discomfort with PAPR. 20% found hearing difficult in PAPR and 30% found speech difficult in EFR |
| Radonovich 2019<sup>50</sup> | 335 | RCT | Filtering facepiece | Simulated | comparison of existing & new devices | R-COMFI score | Probably meaningful improvement with newer devices. Suggests design improvements may lead to better tolerability |
| Rebman 2013<sup>37</sup> | 10 | Observational | Filtering facepiece | Actual | Direct observation | Tolerance time, | 9/10 able to use for 3+ hours before breaks. Approx. 2 violations per hour worked. |
| Fix 2019<sup>51</sup> | 66 | Qualitative | Mixed | - | NA | NA | Complex intersection of personal, social and cultural processes in play |
| Hines 2017<sup>53</sup> | 22 | Qualitative | Mixed | - | NA | NA | Trade-offs between usability (& patient care) vs protection; diversity of opinion on that trade-off; port in a storm rather than the new normal. |
| Hines 2019b<sup>52</sup> | 432 | Cross-sectional survey | Elastomeric facepiece | - | NA | NA | Identified important issues around need for storage of respirator close to patient care in readiness for use and programmes of regular filter replacement and annual fit testing. |

EFR Elastomeric facepiece respirator; FF: Fit factor; FFR: Filtering facepiece respirator; FRSM: fluid resistant surgical mask; NA: not applicable; PAPR: powered air-purifying respirator; RCT: randomised controlled trial; SD: standard deviation; SII: Speech Intelligibility index.

**Table 5.** Summary of review findings.

| Theme | Studies | Participants | Respirators<sup>1</sup> | Main Findings | Strength of evidence for findings |
| --- | --- | --- | --- | --- | --- |
| Need for appropriate fit testing and training | 10 | Fi | FFR (EFR) | At least 10% will need to try more than one respirator in order to achieve fit. Seal check is a poor predictor of fit and is not sufficient. FFR fit markedly diminished in presence of facial hair. | Several large cross-sectional studies with appropriate populations of HCWs |
| Reliability of fit-tested respirator in clinical activity | 7 | 384 | FFR (PAPR) | CPR led to failure of fit in 10-60% of FFR users (3 studies). No failure in PAPR users, no studies with EFR. Generic healthcare activities: 0-30% fit failure with FFR during generic healthcare activities | Consistent finding in small simulation studies. Clinical significance not known. |
| Adherence to standards in practice and effect of training | 3 | 165 | FFR | Failure to follow guidelines for safe use is common both in donning / doffing and during use. Repeated training appears to be necessary to ensure continuing safe respirator fit | Small studies in specific settings. Likely that this is an issue, but unclear how large |
| Impact on clinical performance | 4 | 83 | EFR<br>PAPR<br>(FFR) | Performance of simulated procedures including endotracheal intubation minimally affected. Participants report problems with vision and with hearing. | Small but well conducted simulator studies |
| Impact on clinical communication<sup>2</sup> | 6 | 1741 | EFR<br>PAPR<br>(FFR) | Measured meaningful drop in speech quality (EFR & PAPR) and hearing (PAPR). Subjective identification of difficulties in 20-40% users | Experimental studies indicate meaningful impact likely, surveys vary on perceived extent |
| Impact on comfort | 10 | 2604 | EFR<br>PAPR<br>FFR | Discomfort reported in 15-40% users. Higher with EFR/PAPR than FFR. More than half of users unable to wear for full 8hr shift. | Consistent effect but magnitude highly variable and surveys at high risk of bias |
| HCW & organisation perceptions regarding use | 3 | 1510 | EFR<br>PAPR<br>(FFR) | HCW balance between discomfort and extra protection. Both HCW and organisations indicate important of practical issues (storage, access) and social context of norms and culture. | Two high quality qualitative studies + surveys |

FFR: filtering facepiece respirator; EFR: elastomeric facepiece respirator; PAPR : powered air-purifying respirator.
<sup>1</sup> Respirator types in parentheses appear only infrequently in these studies
<sup>2</sup> Communication comprised 2 direct measurement studies (N=16) and 4 surveys (N=1725)

#### Studies assessing respirator fit

Ten studies assessed respirator fit during static fit-testing or in a series of simple generic manoeuvres (such as speaking, turning or bending at the waist) on healthcare workers. These used either quantitative or qualitative testing, typically after the user had completed a seal check.

Three studies^17-19^ examined simple seal checks by healthcare workers and students. All showed that seal checks prior to formal fit tests are poor predictors of the fit test result. Seal checks gave both false positive and false negative results with positive and negative likelihood ratios both close to 1.^17^ One study found few false negative seal checks but still found that approximately 1 in 4 who passed the seal check failed the fit test and this was unrelated to level of experience.^18^ Together these studies indicate that seal checks without prior fit test are not an appropriate method to assess the efficacy of respirators.

Four studies reported the results of sequential fitting of filtering facepiece respirators until a fit test was passed.^20-23^ In the largest study (N=5024), which used quantitative testing, 82.9% were successfully fitted with the first mask selected by the fitter, 12.3% with the second choice; 4.8% had to try three or more before getting a correct fit.^21^ A second large study (N= 1271), which used qualitative testing, found 87.7% of healthcare workers were successfully fitted with the first choice filtering facepiece respirator. Most, but not all, were successfully fitted with a different one.^20^ A smaller study (N=105) examined the effect of facial hair on fit test and found that the likelihood of successful fit (with a single filtering facepiece respirator type) reduced proportionately to the amount of facial hair present.^22^

#### Studies examining the effect of clinical activities on respirator fit

Seven studies assessed the performance of healthcare workers’ respirators (which had passed initial fit-testing) during simulated clinical activities. Six studies assessed the performance of filtering facepiece respirators and one study assessed powered air-purifying respirators; we identified no studies that had assessed elastomeric facepiece respirators in this way.

Four studies examined the effect on respirator fit of carrying out simulated cardiopulmonary resuscitation chest compressions and one of airway intubation. Three simulated cardiopulmonary resuscitation studies used filtering facepiece respirators and one used powered air-purifying respirators. We report only on participants who had passed a fit test before the simulated activity. In a study of 44 healthcare workers who had passed a fit test with a filtering facepiece respirator, 32 of 44 failed the fit test during at least one of three cycles of chest compression.^24^ In a smaller study which included cardiopulmonary resuscitation as one of a range of nursing activities (N=15), 3 of 15 failed the fit test.^25^ One study (N=45) compared the fit during cardiopulmonary resuscitation of three different filtering facepiece respirators; failure rate varied from 7% to 64%.^26^ One study (N=91) examined the effectiveness of powered air-purifying respirator during cardiopulmonary resuscitation and found that no participant failed the fit test at any stage – a finding which, if replicated, would provide strong support for this kind of mask in CPR contexts.^27^

The simulated intubation study involved emergency physicians (N= 26) using three different types of airway intubation while wearing filtering facepiece respirator after passing a conventional fit test.^28^ 6/24 participants experienced fit failure wearing a cone type of filtering facepiece respirator (though not a folding type) when using direct laryngoscopy compared to none with a video laryngoscope or laryngeal mask airway. This finding is concerning, given the current WHO recommendation that N95 and FFP masks are adequate for this aerosol-generating procedure.

A large study (N=120) of simulated nursing activities, found that 40 of 120 student nurses who had passed a fit test wearing a filtering facepiece respirator failed the fit test during at least one of the activities.^29^ In a smaller but in-depth study with experienced nurses (N=8) who had passed a fit test wearing a filtering facepiece respirator, there were no failures in fit test during a range of clinical activities.^30^

#### Studies of respirator use in practice and the effects of training

Two studies examined fit before and after training and found it improved after training. For healthcare workers with experience of occasional use, training increased fit test pass rates from 15 of 22 to 22 of 22 in one study,^31^ and from 9 of 50 to 20 of 50 in another.^32^ The latter study appears to have tested the effect of training before assessing whether a fit could be obtained with any given respirator. For healthcare worker who had successfully passed a fit test after training, retesting after 3 months (without regular respirator use) found that only 20 of 43 passed a fit test,^33^ suggesting that training needs to be repeated regularly.

Researchers in three studies observed healthcare workers donning and doffing personal protective equipment which included a previously fitted and tested respirator.^34-36^ Non-compliance with recommended technique was observed in approximately half the participants. An intensive observational study following nurses over entire shifts found at least two episodes per hour of touching the respirator during use.^37^

#### Impact of respirator use on clinical performance

Five studies examined the effect of wearing a respirator on performance of skilled clinical tasks. Two crossover studies in which experienced anaesthetic practitioners carried out repeated tracheal intubation in a simulator while wearing elastomeric facepiece respirator, powered air-purifying respirator or neither found a clinically meaningful delay in performance.^38 39^ However, those who wore spectacles reported problems with using these and both respirator types prevented effective chest auscultation to check appropriate tube placement. Two studies examined simulated resuscitation of adults^40^ and children.^41^ Both compared full- and half-face elastomeric facepiece respirators and the paediatric study also included powered air-purifying respirators. There were no statistically significant or clinically meaningful differences in procedure time although several participants reported some impairment of visual field. The study which tested fit of filtering facepiece respirator during intubation showed no adverse effect on performance.^28^

#### Impact of respirator use on communication

Two studies focused on quality of speech communication using a simulated and or real intensive care unit environment. One used human listeners with standardised speech;^13^ the other used an automated approach based on speech sound frequencies ^42^. Both demonstrated that while simple filtering facepiece respirators have only minor effects on speech quality, elastomeric facepiece respirators and to a lesser extent powered air-purifying respirators do impact meaningfully on speech clarity. This corresponds to subjective observations from user surveys in which a negative effect on communication was reported by 20-40% of respondents, with lower satisfaction for elastomeric facepiece and powered air-purifying respirators than filtering facepiece respirators.^43^ A study limited to powered air-purifying respirator users found higher levels of interference with communication, with 60% reporting interference with speaking and 35% reporting interference with hearing.^44^

#### Impact on users

We found one survey which included healthcare workers using one of three different types of respirator: filtering facepiece, elastomeric facepiece and powered air-purifying;^43^ and one survey of powered air-purifying respirator users.^44^ In addition, we found three surveys with more than 100 respondents reporting comfort and usability from filtering facepiece respirators.^45-47^ Two studies particularly focused on headache associated with filtering facepiece respirator use.^48 49^ One study assessed how long clinicians could comfortably wear a respirator through a shift and found that at least half were unable to manage a full 8 hour shift. Filtering facepiece respirators were least well tolerated over a prolonged period; powered air-purifying respirators or filtering facepiece respirators with an expiratory valve were more likely to be tolerated for a long period.^12^ A recent trial compared new respirators with established models and argued that newer designs may reduce discomfort.^50^

One study involved healthcare workers from multiple hospitals in two separate US states and reported data from 1152 respondents (approximately 10% of the invited sample). Of these, 24% used elastomeric facepiece respirators and 23% used powered air-purifying respirators; the remaining 53% regularly used filtering facepiece respirators. Across the different respirator types, rates of perceived discomfort ranged from 15-30%; it was lowest for filtering facepiece respirators and highest for elastomeric facepiece respirators. Approximately 70-80% of healthcare workers reported confidence in the protection afforded by their respirator, with rates being highest in elastomeric facepiece respirator users. The study of powered air-purifying respirator users in cardiopulmonary resuscitation (N=51) reported similar levels of discomfort (39%)^27^.

Studies varied in the way questions were framed and answers reported such that we have not carried out a quantitative synthesis. Nonetheless, the levels reported in these samples appear broadly comparable with the filtering facepiece respirator users in the largest of the studies.^43^

Two studies specifically investigated headache. The first study (N=212) found headaches reported with filtering facepiece respirator use in 37% of healthcare workers with a history of one or more headache disorder and 21% of healthcare workers without prior headache. A second study from the same location found 128/158 nurses reported at least one new headache associated with filtering facepiece respirator use, although three-quarters of these were never more than mild and never required analgesic.

#### Adoption of respirators by healthcare workers and organisations

We found one high quality qualitative study addressing respirator use from a healthcare worker perspective. This study used wide sampling, an evolving analytical strategy and appropriate use of theory^51^ and found that healthcare workers balanced workplace norms and culture against personal and professional judgement and practical issues of access to equipment. A large survey of healthcare workers (N=432) identified substantial logistical issues with the supply, storage and use on-demand of elastomeric facepiece respirators.^52^

We found one large survey of clinical leaders from multiple sites^52^ and one in-depth qualitative study of 11 key informant interviews followed by a healthcare worker focus group.^53^ These identified trade-offs between usability and patient care against protection, with a diversity of opinion on how that trade-off was made. Respondents saw elastomeric facepiece respirators as a temporary defence in unusual circumstances rather than a new normal.

## Discussion

### Statement of principal findings

There are four main findings. First, international standards for respirators apply across different workplace settings and are broadly comparable across jurisdictions. This permits wider choice than the basic disposable filtering facepiece respirators. Second, proper fitting, training in use, and checking at every use are essential for safe respirator use; failures of these are common and result in reduced protection. Third, all respirator types carry a burden to the user of discomfort and interference with communication, which may limit the safe use of respirators for prolonged periods. They appear to have little impact on clinical skills in the short term. Finally, some clinical activities, particularly chest compressions, reduce the protection provided by filtering facepiece respirators.

### Strengths and weaknesses of the study

Strengths of the study was the highly interdisciplinary nature of the team, comprising individuals with expertise in Occupational Medicine (AA), infectious diseases and infection control (X-H C, LR), respirator design and performance (SS) and evidence synthesis (CB, BC, ET and TG); and adherence to Cochrane Rapid Review interim guidance.^7^ This study was a rapid review with a search of two databases, supplemented with hand-searching of references and citations from a sample of high-quality papers^12 21 43 51^ and the personal reference libraries of two of the authors with expertise in the topic (AA and SS). In light of the heterogeneity of studies and reported findings and the need to produce a timely review, we did not carry out a formal analysis of risk of bias. In the context of Covid-19 and related research activity, we recognise that new research is emerging daily and so some of the findings of this review may quickly be superseded.

### Meaning of the study: implications for clinicians and policymakers

Clinicians, particularly those who do not regularly use respiratory protective equipment outside of crises such as Covid-19, need to be aware of the importance of fitting and fit-testing. While the public discourse has mostly centred on the availability of protective equipment, our findings show that professionals’ use of respirators is frequently inadequate. Implementing respirator use requires a system-wide approach which includes availability, fit testing, training, a culture of use and checking, and recognition of the burden that wearing a respirator may add for the busy clinician^54^. During the Covid-19 pandemic there have been some healthcare workers wearing a surgical mask over a fitted facepiece respirator, the reason being to preserve the respirator from direct contamination because of the PPE shortages. This practice may interfere with the face fit of the respirator and impose additional respiratory burden. Where exposure to body fluids is a substantial risk it may be more appropriate to use a reusable respirator (powered air-purifying respirator or elastomeric facepiece respirator) or separate full-face visor with a filtering facepiece respirator. The heterogeneity of healthcare workers face sizes and shapes mean that no single model of filtering facepiece respirator will be suitable for all users; hospitals and other providers and must be prepared to fit users from a range of devices. Hospitals with a substantial level of disposable respirator use should consider whether re-usable respirators (elastomeric facepiece respirator, powered air-purifying respirator) may be both safer for the users and more economical in the long run, particularly if the environmental cost of single use respirators is considered.

### Unanswered questions and future research

We identified two key areas for further research. First there is a need for studies and solutions to the problem of loss of fit in filtering facepiece and elastomeric facepiece respirators during emergency procedures such as chest compression (either these products need modifying or the guidance needs to specifically recommend the higher-grade powered air purifying respirators. Second, designers and manufacturers should work to develop respirator designs which reduce user discomfort and minimise disruption of communication for respirator users.

## Conclusion

A wide range of respirator types and models can be used in patient care during the Covid-19 pandemic. Careful consideration of performance and impact of respirators is needed to maximise protection of healthcare workers and minimise disruption to the delivery of care.

## Data Availability

This is a secondary analysis of published primary data.

## APPENDIX 1. SEARCH STRATEGY

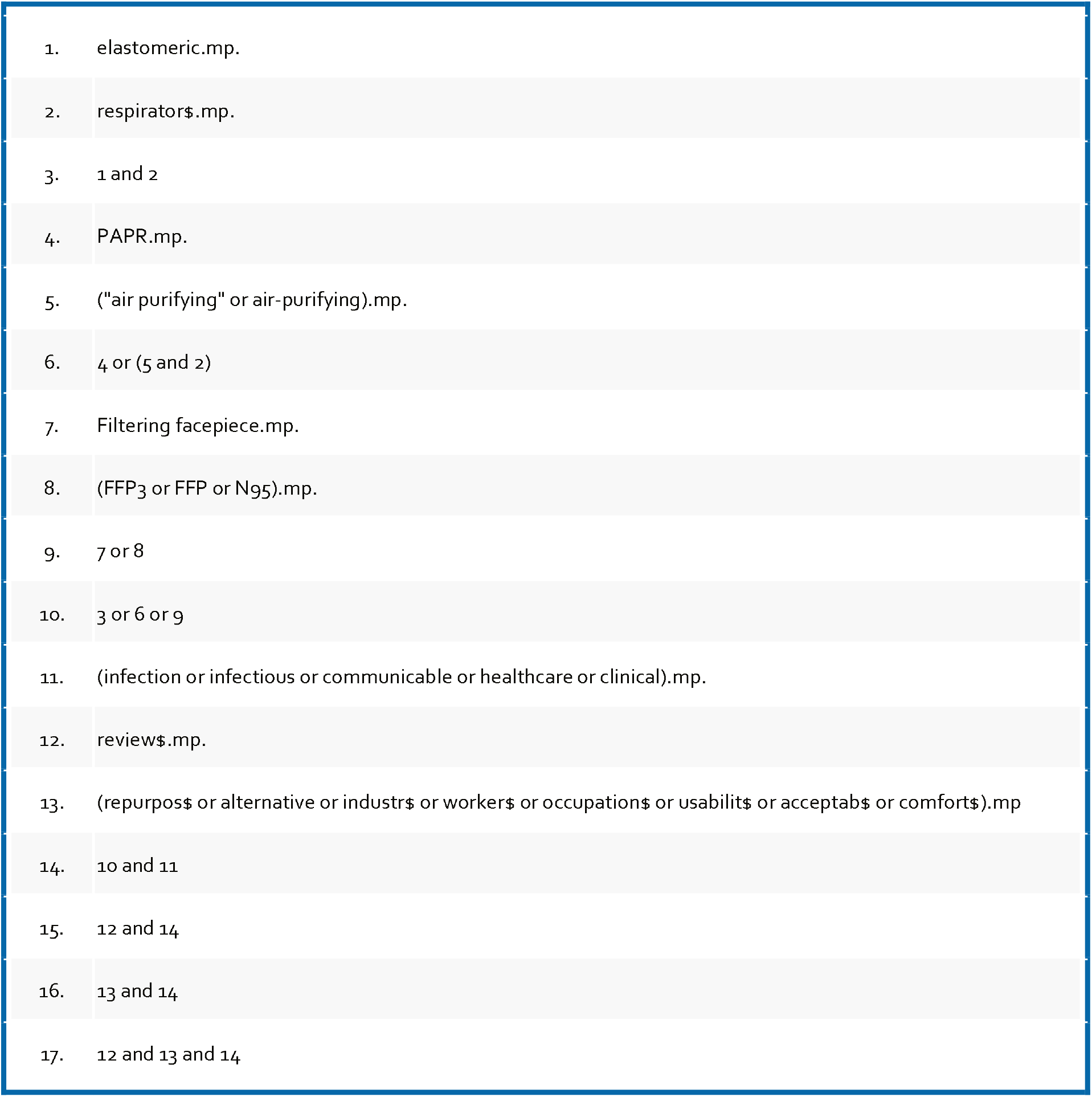

## Contributorship and guarantor

The article was collaboratively developed as part of a wider series of evidence reviews on personal protective equipment edited by TG and overseen by the Oxford Covid-19 Evidence Review Service. SS conceptualised the review and undertook extensive background desk research on respirator standards. CB led the shaping of the methodology to align with formal systematic review guidance. CB &SS undertook searches. CB & BC contributed to data extraction. CB led the synthesis. AA provided specialist occupational medicine expertise. X-HC and LR provided specialist infection control expertise. CB wrote the first draft of the paper, to which all authors made contributions. All authors approved the final manuscript. CB is corresponding author and guarantor.

## Acknowledgements

None

## How patients were involved in the creation of this article

Members of the public were not involved in the review or the writing of the paper.

## References

1. England PH. Considerations for acute personal protective equipment (PPE) shortages 2020 [updated 3/5/2020. Available from: https://www.gov.uk/government/publications/wuhan-novel-coronavirus-infection-prevention-and-control/managing-shortages-in-personal-protective-equipment-ppe accessed 15/5/2020.

2. Prevention CfdCa. Strategies for Optimizing the Supply of N95 Respirators 2020 [updated 22 April 2020. Available from: https://www.cdc.gov/coronavirus/2019-ncov/hcp/respirators-strategy/index.html accessed 15/5/2020.

3. Kamerow D. Covid-19: the crisis of personal protective equipment in the US. BMJ 2020;369:m1367. doi: 10.1136/bmj.m1367

4. Friese CR, Veenema TG, Johnson JS, et al. Respiratory Protection Considerations for Healthcare Workers During the COVID-19 Pandemic. Health Secur 2020 doi: 10.1089/hs.2020.0036 [published Online First: 2020/04/23]

5. Verbeek JH, Rajamaki B, Ijaz S, et al. Personal protective equipment for preventing highly infectious diseases due to exposure to contaminated body fluids in healthcare staff. Cochrane Database of Systematic Reviews 2020(4) doi: 10.1002/14651858.CD011621.pub4

6. Poller B, Tunbridge A, Hall S, et al. A unified personal protective equipment ensemble for clinical response to possible high consequence infectious diseases: A consensus document on behalf of the HCID programme. J Infect 2018;77(6):496–502. doi: 10.1016/j.jinf.2018.08.016 [published Online First: 2018/09/04]

7. Garritty C, Gartlehner G, Kamel C, et al. Cochrane Rapid Reviews. Interim Guidance fromthe Cochrane Rapid Reviews Methods Group, 2020.

8. Burton C. Respirator selection and use review, protocol 1.0: Open Science Framework; 2020 [Available from: https://osf.io/a4ym3/ accessed 13/5/2020.

9. Greenhalgh T, Peacock R. Effectiveness and efficiency of search methods in systematic reviews of complex evidence: audit of primary sources. Bmj 2005;331(7524): 1064–5. doi: 10.1136/bmj.38636.593461.68 [published Online First: 2005/10/19]

10. Cooke A, Smith D, Booth A. Beyond PICO: the SPIDER tool for qualitative evidence synthesis. Qual Health Res 2012;22(10):1435–43. doi: 10.1177/1049732312452938 [published Online First: 2012/07/26]

11. Medicine Io. Reusability of Facemasks During an Influenza Pandemic: Facing the Flu Washington, DC, 2006.

12. Radonovich LJ, Cheng J, Shenal BV, et al. Respirator Tolerance in Health Care Workers. JAMA 2009;301(1):36–38. doi: 10.1001/jama.2008.894

13. Radonovich LJ, Yanke R, Cheng J, et al. Diminished speech intelligibility associated with certain types of respirators worn by healthcare workers. Journal of Occupational and Environmental Hygiene 2010;7(1):63–70. doi: 10.1080/15459620903404803

14. Toomey ES, M.; Conway, Y.; Devane, D.; Burton, C.; Jackson, T.; Smith, S.; Straube, S.; Adisesh, A.; Durand-Moreau, Q.; Chen, XH.; Ross, L.; Greenhalgh, T.. Protocol: Overview of recommendations and evidence for reuse and/or extended use of respiratory protective equipment (RPE) for the prevention of COVID-19. 2020. https://osf.io/8qxr7/.

15. Shaffer RE, Janssen LL. Selecting models for a respiratory protection program: What can we learn from the scientific literature? American Journal of Infection Control 2015;43(2): 127–32. doi: 10.1016/j.ajic.2014.10.021

16. National Academies of Sciences E, Medicine. Reusable Elastomeric Respirators in Health Care: Considerations for Routine and Surge Use. Washington, DC: The National Academies Press 2019.

17. Lam SC, Lui AKF, Lee LYK, et al. Evaluation of the user seal check on gross leakage detection of 3 different designs of N95 filtering facepiece respirators. American journal of infection control 2016;44(5):579–86. doi: https://dx.doi.org/10.1016/j.ajic.2015.12.013

18. Danyluk Q, Hon C-Y, Neudorf M, et al. Health Care Workers and Respiratory Protection: Is the User Seal Check a Surrogate for Respirator Fit-Testing? Journal of Occupational and Environmental Hygiene 2011;8(5):267–70. doi: 10.1080/15459624.2011.566016

19. Derrick JL, Chan YF, Gomersall CD, et al. Predictive value of the user seal check in determining half-face respirator fit. The Journal of hospital infection 2005;59(2):152–5.

20. McMahon E, Wada K, Dufresne A. Implementing fit testing for N95 filtering facepiece respirators: practical information from a large cohort of hospital workers. American journal of infection control 2008;36(4):298–300. doi: https://dx.doi.org/10.1016/j.ajic.2007.10.014

21. Wilkinson IJ, Pisaniello D, Ahmad J, et al. Evaluation of a large-scale quantitative respirator-fit testing program for healthcare workers: survey results. Infection control and hospital epidemiology 2010;31(9):918–25. doi: https://dx.doi.org/10.1086/655460

22. Sandaradura I, Goeman E, Pontivivo G, et al. A close shave? Performance of P2/N95 respirators in healthcare workers with facial hair: results of the BEARDS (BEnchmarking Adequate Respiratory DefenceS) study. The Journal of hospital infection 2020 doi: https://dx.doi.org/10.1016/j.jhin.2020.01.006

23. Lee S, Kim H, Lim T, et al. Simulated workplace protection factors for respirators with N95 or higher filters for health care providers in an emergency medical centre: A randomized crossover study. Hong Kong Journal of Emergency Medicine 2017;24(6):282–89. doi: http://dx.doi.org/10.1177/1024907917735088

24. Hwang SY, Yoon H, Yoon A, et al. N95 filtering facepiece respirators do not reliably afford respiratory protection during chest compression: A simulation study. The American journal of emergency medicine 2020;38(1): 12–17. doi: https://dx.doi.org/10.1016/j.ajem.2019.03.041

25. Sietsema M, Brosseau LM. Are quantitative fit factors predictive of respirator fit during simulated healthcare activities? Journal of occupational and environmental hygiene 2018;15(12):803–09. doi: https://dx.doi.org/10.1080/15459624.2018.1515490

26. Shin H, Oh J, Lim TH, et al. Comparing the protective performances of 3 types of N95 filtering facepiece respirators during chest compressions: A randomized simulation study. Medicine 2017;96(42):e8308. doi: https://dx.doi.org/10.1097/MD.0000000000008308

27. Park SH, Hwang SY, Lee G, et al. Are loose-fitting powered air-purifying respirators safe during chest compression? A simulation study. The American journal of emergency medicine 2020 doi: https://dx.doi.org/10.1016/j.ajem.2020.03.054

28. Kang H, Lee Y, Lee S, et al. Protection afforded by respirators when performing endotracheal intubation using a direct laryngoscope, GlideScope R, and i-gel R device: A randomized trial. PloS one 2018;13(4):e0195745. doi: https://dx.doi.org/10.1371/journal.pone.0195745

29. Suen LKP, Yang L, Ho SSK, et al. Reliability of N95 respirators for respiratory protection before, during, and after nursing procedures. American journal of infection control 2017;45(9):974–78. doi: https://dx.doi.org/10.1016/j.ajic.2017.03.028

30. Hauge J, Roe M, Brosseau LM, et al. Real-time fit of a respirator during simulated health care tasks. Journal of Occupational and Environmental Hygiene 2012;9(10):563–71. doi: 10.1080/15459624.2012.711699

31. Kim H, Lee J, Lee S, et al. Comparison of fit factors among healthcare providers working in the Emergency Department Center before and after training with three types of N95 and higher filter respirators. Medicine 2019;98(6):e14250. doi: https://dx.doi.org/10.1097/MD.0000000000014250

32. Winter S, Thomas JH, Stephens DP, et al. Particulate face masks for protection against airborne pathogens - one size does not fit all: an observational study. Critical care and resuscitation: journal of the Australasian Academy of Critical Care Medicine 2010;12(1):24–7.

33. Lee MC, Takaya S, Long R, et al. Respirator-fit testing: does it ensure the protection of healthcare workers against respirable particles carrying pathogens? Infection control and hospital epidemiology 2008;29(12): 1149–56. doi: https://dx.doi.org/10.1086/591860

34. Beam EL, Hotchkiss EL, Gibbs SG, et al. Observed variation in N95 respirator use by nurses demonstrating isolation care. American journal of infection control 2018;46(5):579–80. doi: https://dx.doi.org/10.1016/j.ajic.2017.11.019

35. Nichol K, McGeer A, Bigelow P, et al. Behind the mask: Determinants of nurse’s adherence to facial protective equipment. American journal of infection control 2013;41(1):8–13. doi: https://dx.doi.org/10.1016/j.ajic.2011.12.018

36. Mumma JM, Durso FT, Casanova LM, et al. Common Behaviors and Faults When Doffing Personal Protective Equipment for Patients With Serious Communicable Diseases. Clinical infectious diseases: an official publication of the Infectious Diseases Society of America 2019;69(Supplement_3):S214-S20. doi: https://dx.doi.org/10.1093/cid/ciz614

37. Rebmann T, Carrico R, Wang J. Physiologic and other effects and compliance with long-term respirator use among medical intensive care unit nurses. American Journal of Infection Control 2013;41(12):1218–23. doi: 10.1016/j.ajic.2013.02.017

38. Candiotti KA, Rodriguez Y, Shekhter I, et al. A comparison of different types of hazardous material respirators available to anesthesiologists. American journal of disaster medicine 2012;7(4):313–9. doi: https://dx.doi.org/10.5055/ajdm.2012.0104

39. Schumacher J, Arlidge J, Dudley D, et al. The impact of respiratory protective equipment on difficult airway management: a randomised, crossover, simulation study. Anaesthesia 2020 doi: 10.1111/anae.15102

40. Schumacher J, Runte J, Brinker A, et al. Respiratory protection during high-fidelity simulated resuscitation of casualties contaminated with chemical warfare agents. Anaesthesia 2008;63(6):593–8. doi: https://dx.doi.org/10.1111/j.1365-2044.2008.05450.x

41. Schumacher J, Gray SA, Michel S, et al. Respiratory protection during simulated emergency pediatric life support: a randomized, controlled, crossover study. Prehospital and disaster medicine 2013;?(1):33–8. doi: https://dx.doi.org/10.1017/S1049023X12001525

42. Palmiero AJ, Symons D, Morgan JW, 3rd, et al. Speech intelligibility assessment of protective facemasks and air-purifying respirators. Journal of occupational and environmental hygiene 2016;13(12):960–68.

43. Hines SE, Brown C, Oliver M, et al. User acceptance of reusable respirators in health care. American Journal of Infection Control 2019;47(6):648–55. doi: 10.1016/j.ajic.2018.11.021

44. Khoo K-L, Leng P-H, Ibrahim IB, et al. The changing face of healthcare worker perceptions on powered air-purifying respirators during the SARS outbreak. Respirology (Carlton, Vic) 2005; 10(1):107–10.

45. Baig AS, Knapp C, Eagan AE, et al. Health care workers’ views about respirator use and features that should be included in the next generation of respirators. American journal of infection control 2010;38(1): 18–25. doi: https://dx.doi.org/10.1016/j.ajic.2009.09.005

46. Brosseau LM, Conroy LM, Sietsema M, et al. Evaluation of Minnesota and Illinois hospital respiratory protection programs and health care worker respirator use. Journal of occupational and environmental hygiene 2015; 12(1): 1 -15. doi: https://dx.doi.org/10.1080/15459624.2014.930560

47. Bryce E, Forrester L, Scharf S, et al. What do healthcare workers think? A survey of facial protection equipment user preferences. The Journal of hospital infection 2008;68(3):241–7. doi: https://dx.doi.org/10.1016/j.jhin.2007.12.007

48. Lim ECH, Seet RCS, Lee KH, et al. Headaches and the N95 face-mask amongst healthcare providers. Acta neurologica Scandinavica 2006;113(3):199–202.

49. Ong JJY, Bharatendu C, Goh Y, et al. Headaches Associated With Personal Protective Equipment - A Cross-Sectional Study Among Frontline Healthcare Workers During COVID-19. Headache 2020 doi: https://dx.doi.org/10.1111/head.13811

50. Radonovich LJ, Wizner K, LaVela SL, et al. A tolerability assessment of new respiratory protective devices developed for health care personnel: A randomized simulated clinical study. PloS one 2019;14(1):e0209559. doi: https://dx.doi.org/10.1371/journal.pone.0209559

51. Fix GM, Reisinger HS, Etchin A, et al. Health care workers’ perceptions and reported use of respiratory protective equipment: A qualitative analysis. American Journal of Infection Control 2019;47(10):1162–66. doi: 10.1016/j.ajic.2019.04.174

52. Hines SE, Brown C, Oliver M, et al. Storage and Availability of Elastomeric Respirators in Health Care. Health security 2019;17(5):384–92. doi: https://dx.doi.org/10.1089/hs.2019.0039

53. Hines SE, Mueller N, Oliver M, et al. Qualitative Analysis of Origins and Evolution of an Elastomeric Respirator-based Hospital Respiratory Protection Program. Journal of the International Society for Respiratory Protection 2017;34(2):95–110.

54. Houghton C, Meskell P, Delaney H, et al. Barriers and facilitators to healthcare workers’ adherence with infection prevention and control (IPC) guidelines for respiratory infectious diseases: a rapid qualitative evidence synthesis. Cochrane Database Syst Rev 2020;4(4):Cd013582. doi: 10.1002/14651858.Cd013582 [published Online First: 2020/04/22]

